# Examining the Influence of the Budget Execution Processes on the Efficiency of County Health Systems in Kenya

**DOI:** 10.1101/2022.07.26.22277737

**Authors:** Anita Musiega, Benjamin Tsofa, Lizah Nyawira, Rebecca G Njuguna, Joshua Munywoki, Kara Hanson, Andrew Mulwa, Sassy Molyneux, Isabel Maina, Charles Normand, Julie Jemutai, Edwine Barasa

**Author notes:** **First and Corresponding Author:** Anita Musiega, **Address:** P.O Box 43640-00100 Nairobi, Kenya, **Email address:**.

## Abstract

Public Financial Management (PFM) processes are a driver of health system efficiency. The budget execution process is the stage in the PFM cycle where health system inputs are translated into outputs and outcomes. This study examined how the budget execution process influenced the efficiency of county health systems in Kenya. We conducted a concurrent mixed methods case study using counties classified as relatively efficient (n=2) and relatively inefficient (n=2) in a related quantitative analysis as our cases. We developed a conceptual framework from a literature review to guide the development of tools and analysis. We collected qualitative data through document reviews, and in-depth interviews (n=70) with actors from health and finance sectors at the national and county level. We collected quantitative data from secondary sources, including budgets and budget reports. We analyzed qualitative data using the thematic approach and carried out descriptive analyses on quantitative data. The budget execution processes within counties in Kenya were characterized by poor budget credibility, cash disbursement delays, limited provider autonomy, and poor procurement practices. These challenges were linked to an inappropriate input mix that compromised the capacity of county health systems to deliver healthcare services, misalignment between county health needs and the use of resources, reduced staff motivation and productivity, procurement inefficiencies, and reduced county accountability for finances and performance. The efficiency of county health systems in Kenya can be enhanced by improving budget credibility, cash disbursement processes, procurement processes and improved provider autonomy.

## INTRODUCTION

Achieving Universal Health Coverage (UHC) goals depends on allocating sufficient resources to health and using the allocated resources efficiently (Barroy, Sparkes and Dale, 2016). There are two types of efficiency, allocative efficiency which entails having the best input combination for output maximization (Schick, 1999) and technical efficiency which means getting maximum outputs for available inputs or using the least possible inputs for a given set of outcomes (Schick, 1999). Inefficiencies within the health sector results in wastage of 20-40% of health resources (World Health Organization, 2010). Improving efficiency is an important source of increased health resources (Barroy, Sparkes and Dale, 2016; Barroy *et al*., 2018).

Several studies have identified Public Financial Management (PFM) processes as a driver of health system efficiency (Piatti-Fünfkirchen and Schneider, 2018a; Zeng *et al*., 2021). Public health resources are allocated, used, and accounted for using the PFM processes. PFM happens within the budget cycle – budget formulation and approval, execution, and evaluation.

This paper focuses on the relationship between budget execution and efficiency of county health systems. Budget execution encompasses the provision of promised revenues and the use of these resources to achieve health system objectives (Piatti-Fünfkirchen and Schneider, 2018b). While there are challenges across all the three aspects of the budget process in LMICs, the problems are worse downstream (at budget execution and evaluation) (World Health Organization (WHO), 2016; Barroy *et al*., 2019). Government budgets are better made than they are executed (World Health Organisation, 2018). While it is difficult to execute a poorly formulated budget, it is possible to execute a well formulated budget (Piatti-Fünfkirchen *et al*., 2021). Well formulated budgets that incorporate society views and health system needs are of no value if they are not implemented (Piatti-Fünfkirchen *et al*., 2021). Several countries have reported challenges with their budget execution processes including poor budget credibility, poor budget absorption, corruption, and misappropriation of funds (Asante, Zwi and Ho, 2006; G Gwati Ministry of Health and Child Care and Training and Research Support Centre, 2015; Piatti-Fünfkirchen and Schneider, 2018a). All these challenges can likely compromise the efficiency of the health sector (Piatti-Fünfkirchen *et al*., 2021).

Kenya has a devolved system of governance with a national government and 47 county governments (Government of Kenya, 2010). Within the health sector, the national government has policy and regulatory functions, while county governments have service provision functions (Tsofa *et al*., 2017). County governments are funded through a funding allocation from the national government and locally generated funds. Counties are responsible for allocation and implementation of budgets at their level (Tsofa *et al*., 2017). Like other low- and middle-income countries (LMICs), Kenya has reported challenges in the budget execution process that can likely compromise efficiency. For example, at the national level, one study reported poor budget absorption, poor procurement practices and cash disbursement delays (Glenngård and Maina, 2007). At the county level, studies have reported reduced provider autonomy (Barasa *et al*., 2017), inadequate budget allocation (Mbau *et al*., 2018) and ad hoc reallocation of health resources during execution (Waithaka *et al*., 2018). Understanding how the budget execution structures and processes interact to influence the efficiency of the health system is an important research question.

This study is part of a larger, phased study that examined the efficiency of county health systems in Kenya (the Kenya Efficiency Study). Phase one of the study examined and reported stakeholder perceptions about the factors that affect the efficiency of county health systems in Kenya and identified PFM as one of those factors (Nyawira *et al*., 2020). Phase two measured the level and determinants of county health system efficiency. This phase ranked the 47 counties using an efficiency score and identified the absorption of county budgets (a budget execution issue) as a determinant of county health system technical efficiency (Barasa *et al*., 2021). This paper reports part of the findings of the third and final phase of the study, which entailed in-depth case studies in selected counties to examine identified determinants of county health system efficiency. Specifically, in this study we examine how the budget execution process influences the efficiency of county health systems in Kenya.

## METHODS

### Conceptual Framework

We developed a conceptual framework (Figure 1) from a scoping review on the effects of budget execution processes on the efficiency of health systems. We found five potential dimensions of the budget execution process that may influence the efficiency of health systems: budget credibility, cash disbursement processes, procurement processes, provider autonomy, and financial management information systems. These dimensions may influence health system efficiency directly or through influencing the budget formulation and evaluation processes. This conceptual framework guided the development of tools and data analysis.

**Figure 1:**
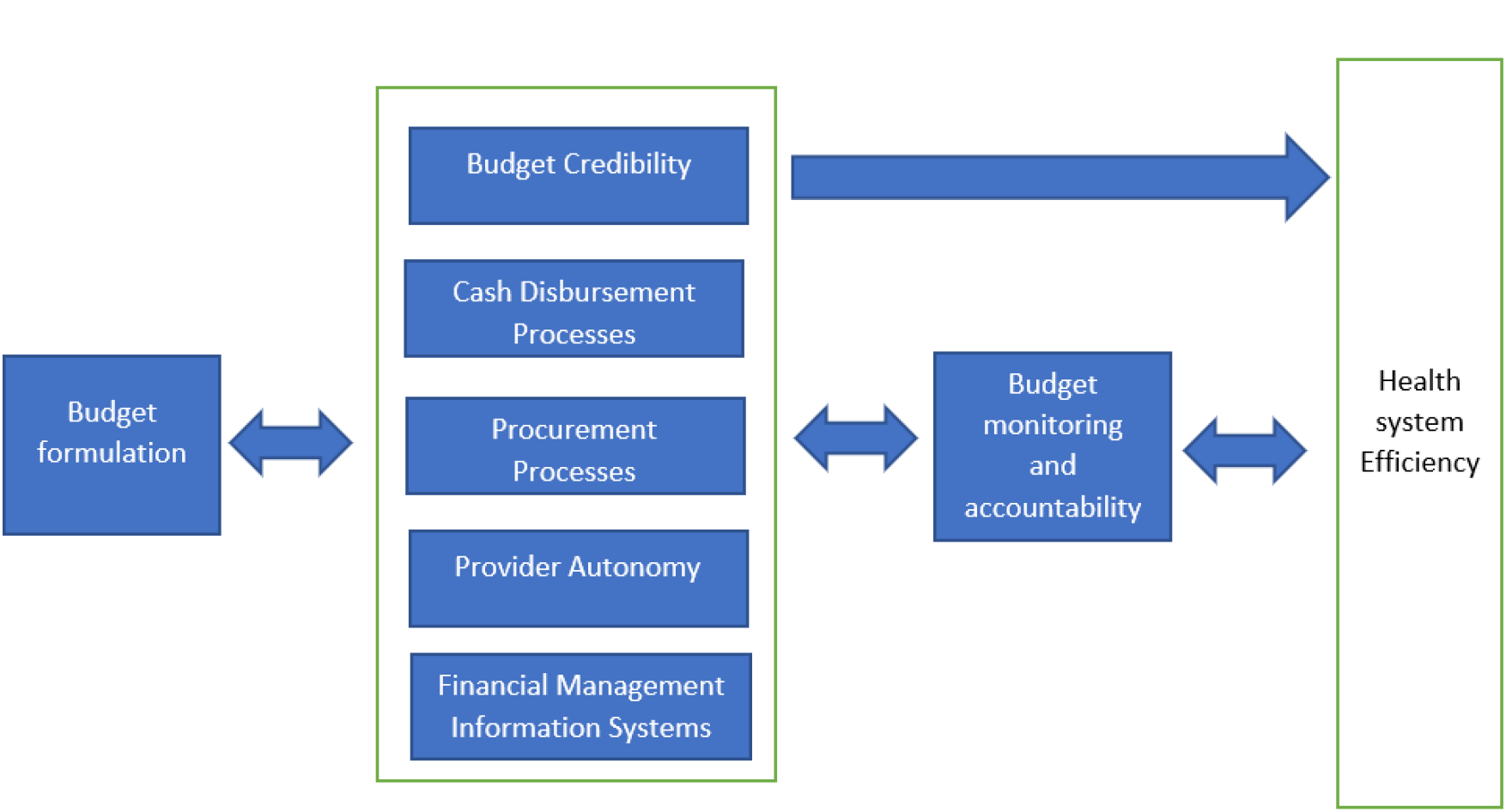
Conceptual Framework

**Figure 2:**
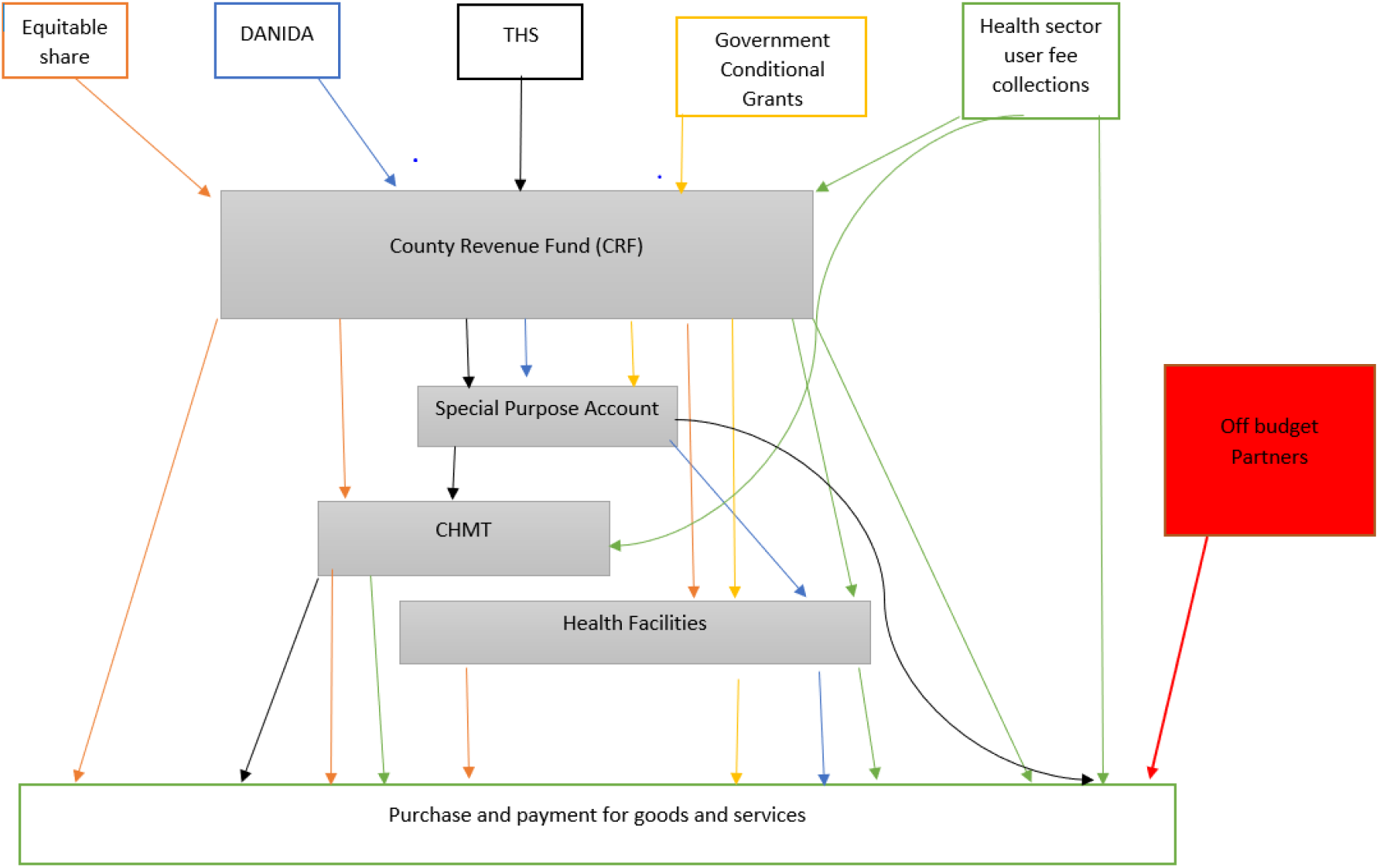
Budget Execution Process within county health systems in Kenya

### Study Design

We conducted a mixed methods case study with data collected through in-depth interviews and document reviews. The case studies allowed for the exploration of the budget execution process within counties in Kenya. We used qualitative methods to explore stakeholder perceptions and quantitative methods to analyse budget data to contextualize the qualitative data.

### Study Cases

We selected four cases - counties - using the level of technical efficiency reported in phase two of the Kenya Efficiency Study (Barasa *et al*., 2021). We then took into consideration other county characteristics that were identified by the quantitative efficiency analysis as determinants of efficiency. These are population size and the prevalence of HIV/AIDs. That is, we selected counties with varying population size and HIV prevalence. We selected two counties with efficiency scores above 0.9 (A and B) as the efficient counties and two counties with efficiency scores below 0.5 (C and D) as the inefficient counties (Table 1).

**Table 1:** County profiles

| County | Efficiency Score* | Population (2019) | Total County Public Health Recurrent Expenditure (2018/2019) KES | Total County Public Health Development Expenditure (2018/2019) KES | Per Capita County Public Health Expenditure (2018/2019) KES | Percentage of public facilities | Percentage of private facilities |
| --- | --- | --- | --- | --- | --- | --- | --- |
| A | 0.9 | 1,163,186 | 1,884,620,000.00 | 46490000.00 | 1660.190202 | 62% | 38% |
| B | 0.9 | 990,341 | 949,629,480 | 615,371,170 | 1580.264424 | 47% | 53% |
| C | 0.4 | 1,131,950 | 2,121,046,189 | 370,754,248 | 2201.334367 | 63% | 37% |
| D | 0.5 | 315,943 | 749,054,078 | 167,564,044 | 2901.21358 | 71% | 29% |

### Data Collection

We collected data using in-depth interviews and document reviews. We selected 70 participants (Table 2) from health and finance departments as both were involved in the budget execution process. The participants were selected from the national level, counties, and development partners. The development of topic guides was guided by the study’s conceptual framework. Interviews were audio recorded and took between 45-90 minutes. Study respondents provided signed informed consent. We extracted budget data from a review of relevant documents (Table 3).

**Table 2:** Study Participants Profile

|  |  |  |
| --- | --- | --- |
|  | County-Level Respondents | National Level Respondents |

| Interviewee group | A | B | C | D |  |
| --- | --- | --- | --- | --- | --- |
| Health Managers | 4 | 6 | 3 | 3 | 1 |
| Finance Managers | 4 | 1 | 2 | 5 | 1 |
| Sub County Health Managers | 0 | 3 | 2 | 0 | - |
| Facility Health Managers | 7 | 9 | 5 | 9 | - |
| Donors | - | - | - | - | 6 |
| Sub totals | 15 | 19 | 12 | 17 | 8 |
| Total | **70** |  |  |  |  |

**Table 3:** Documents reviewed

| Documents Type | Total Reviewed |
| --- | --- |
| County Votebooks 17/18 | 3 |
| County Votebooks 18/19 | 3 |
| County Fiscal Strategy Paper 18/19 | 4 |
| County Budget Review and Outlook Paper 18/19 | 1 |
| County Budget Operationalization Manual | 1 |
| Public Finance Management Act | 1 |
| Public Finance Management Guidelines | 1 |
| County Governments Budget Implementation Review Reports | 1 |
| County Audit Reports FY 18/19 | 4 |
| Total | 19 |

### Data Analysis

We transcribed all the recordings and transferred the data to NVIVO for analysis. We analyzed the data using a thematic approach that entailed developing a pre -analysis theme followed by identification, organization, description, analysis, and reporting of themes found in a data set (Braun and Clarke, 2006). We conducted descriptive analysis on budget data on Microsoft excel. We used quantitative data to contextualize the results of the qualitative data.

## RESULTS

In this section we first present the budget execution process within county health systems in Kenya then we present the following five dimensions of the study’s conceptual framework (Figure 1): 1) Budget credibility 2) Cash disbursement process 3) Procurement process 4) Provider autonomy and 5) Financial Management Information System. Summary findings per county are outlined in table 4.

**Table 4:** Summary findings per county

| Issue | County A | County B | County C | County D |
| --- | --- | --- | --- | --- |
| Honoring of budget releases (from national treasury) | Fully honored | Fully honored | Fully honored | Fully honored |
| Budget implemented as per plans developed | Executed as per plans | Not executed as planned | Not executed as planned | Not executed as planned |
| Timely payment for goods and services | Timely | Delayed | Delayed | Delayed |
| Competitive bidding for tenders | Competitive | Not competitive | Not competitive | Not competitive |
| Procurement process delivers goods and services timely | Timely | Delayed | Delayed | Delayed |
| CDOH gets value for money from procured goods and services | Value for money | No value for money | No value for money | No value for money |
| Frontline workers involved in procurement | Partially involved | Not involved | Partially involved | Not involved |
| Provider financial autonomy over own source revenue | Partial autonomy | Partial autonomy (until October 2020) | No autonomy | No autonomy |
| Access to IFMIS<br>granted to the<br>CDOH | Departmental<br>access | Departmental<br>access | Departmental<br>access | No departmental<br>access |

### Budget Execution Process

There was no standard approach to executing health budgets within county health systems in Kenya. The county government received revenue from multiple sources and used the revenue through multiple channels. There were five revenue sources; equitable share allocation by the national government, on budget donor conditional grants from the Danish Development Aid Agency (DANIDA) and the World Bank’s Transforming Health Systems project (THS), government conditional grants (User fee forgone and tertiary hospitals (Level 5) grants), own source revenue (user fees collected and insurance reimbursements) and off budget partner support. Expenditure of the collected revenue also took place at multiple levels including county treasury (the county revenue fund - CRF and special purpose account - SPA), county health management team (CHMT), health facilities and at partner level by off budget partners.

### Budget Credibility

Budget credibility influenced county health system efficiency through 1) timely realization of expected revenue for budget execution, 2) the implementation of the budget as per plans developed during formulation.

### Despite counties receiving their full equitable share, they did not always disburse budgets allocated to county departments of health

CDOH respondents linked the failure to honor approved budgets to 1) county failure to realize revenue targets from some sources (Table 5) 2) limited transparency in the budget execution process 3) minimal departmental control over resources and 4) failure to meet conditions attached to conditional grants:

> *“we don’t get money as a chunk from the national treasury it trickles in periodically. It is then the department of finance that decides how much to give to health from that disbursement. So budgeting is one thing, but executing that budget in the context of the counties is very difficult” County Health Manager County B*

**Table 5:** Actual County budget receipts as a percentage of Budget Allocation

| Source of fund | Percentage released |  |  |  |  |  |  |  |
| --- | --- | --- | --- | --- | --- | --- | --- | --- |
|  | County A |  | County B |  | County C |  | County D |  |
|  | 2017/2018 | 2018/2019 | 2017/2018 | 2018/2019 | 2017/2018 | 2018/2019 | 2017/2018 | 2018/2019 |
| Equitable share | 100% | 99.83% | 100% | 100% | 100% | 100% | 100% | 100% |
| Own Source revenue | 84.39% | 76.57% | 61.51% | 149% | 90.12% | 58% | 145.70% | 106% |
| THS | 31.20% | 47.30% | 31.30% | 40.90% | 111% | 48.10% | 45.50% | 51.10% |
| DANIDA Grant | 47.00% | 143.10% | 64.50% | 100% | 100% | 100% | 100% | 100% |
| User fee forgone grant | 106.40% | 100% | 94.90% | 100% | 51% | 100% | 50.20% | 100% |

Source: County Governments Annual Budget Implementation Review Reports (2017/2018, 2018/2019)

Failure to receive disbursements of expected revenue influenced efficiency in various ways. First, CDoHs had to forfeit key budget items. This compromised the health system input mix and compromised the capacity of facilities to provide health services.

> *“We included the repair of our faulty solar system in the budget. However, it was not funded. During blackouts, we are forced to conduct maternal deliveries in darkness. The mothers leave the hospital with a negative perception. They discourage other mothers from coming to the hospital. We put a lot of effort in mobilizing mothers to deliver at the hospital but they don’t come because we have challenges” Facility Manager County D*

Second, failure to realize expected revenue resulted in late payment and nonpayment of bills from private suppliers of health commodities (Table 6). This led to private suppliers subsequently refusing to supply health commodities the counties.

**Table 6:** Summary of Total County Pending Bills (KES Millions)

|  | 2017/2018 |  |  | 2018/2019 |  |  |
| --- | --- | --- | --- | --- | --- | --- |
|  | Development | Recurrent | Total | Development | Recurrent | Total |
| County A | 84.15 | 235.2 | 319.35 | 51.73 | 229.8 | 281.53 |
| County B | 313.3 | 468.28 | 781.58 | 421.71 | 317.51 | 739.22 |
| County C | 666.21 | 80.27 | 746.48 | 949.74 | 89.7 | 1039.44 |
| County D | 853.44 | 95.59 | 949.03 | 891.6 | 490.51 | 1382.11 |
| National Total | 28055.38 | 80356.05 | 108411.43 | 11626.34 | 22911.73 | 34538.07 |
| National Average | 596.92 | 1709.70 | 2306.63 | 247.37 | 487.48 | 734.85 |

Source: Controller of Budget

> *“Our Local Service Order (LSO) are never honoured because disbursements were not honoured. Yet the goods/services were delivered. This results in debts. Because of this, some of our suppliers decline to furnish our orders*.*” Facility health Manager County B*

Third, it compromised the ability of the CDOH management to hold health facility managers accountable for performance. When disbursements to health facilities were not honored, the CDOH supervisors lacked the legitimacy to supervise to ensure delivery of services.

> *“Supervision is very weak in health. The CHMT is embarrassed to visit health facilities to supervise what they’ve not funded*. Besides i*f someone came here to supervise me, I would be very reluctant, and I believe any MOH (Medical Officer of Health) would be very reluctant to be supervised on what has not been funded despite budgeting and approval” Facility Health Manager County C*

### The budget was not implemented as per the plans developed during budget formulation

In three of the four study counties, utilization of funds deviated from the formulated budgets and plans. The CDOH respondents noted that the COB approved expenditure based on requests that were directly linked to approved budgets. However, once these funds were availed, they were used at the discretion of the county treasury.

Respondents noted that budget execution that deviated from existing budgets and plans without a clear need for reallocation influenced efficiency in several ways. First, expenditure deviated from the county needs, compromising the achievement of health system targets.

> *“One can easily come up with a plan which is neither in the budget nor in the AWP. Our plans are made to achieve targeted indicators. So, if you conduct an activity that is outside your plan, you may not be working towards the intended goal” County Health Manager County D*

Second, unapproved deviation from existing plans and budgets limited accountability. Departments spending the money were unable to track expenditure and verify service delivery.

### Cash Disbursement Processes

CDOH and treasury respondents in county A noted that their cash disbursement processes were well organized and timely, respondents from the other three study counties noted that their processes were bureaucratic and late. The challenges with cash disbursement processes influenced efficiency by influencing 1) timeliness of payment and 2) how payments were prioritized.

### In three of the four counties (B, C, D), cash disbursements were late

Lateness in disbursement of finances hindered efficiency in several ways. First, it compromised service delivery and hence potentially negatively affected health outcomes.

> *“Our performance depends on the flow of funds. As the CEC I have a performance contract that should be executed within specified timelines*… *But the flow of funds has been the greatest challenge. Because if funds don’t flow, you may plan but you can’t implement” County Health Manager County B*

Second, some disbursements came too late in the financial year influencing the ability to honor commitments for already incurred expenses (Table 7), absorption for un-incurred expenses (Table 4) and ultimately resulting in pending bills (Table 8).

**Table 7:** County Department of Health Total Outstanding Commitments at the end of FY 18/19

| Details | County A | County B | County C | County D |
| --- | --- | --- | --- | --- |
| Recurrent outstanding commitments | 15,627,958.00 | 11,920,166.00 | 0 | 4,888,500.00 |
| Development outstanding commitments | 37,629,926.00 | 998,130.00 | 161,442,285.00 | 519,535.00 |
| Total outstanding commitments | 53,257,884.00 | 12,918,296.00 | 161,442,285.00 | 5,408,035.00 |
| Outstanding commitments as a percentage of total commitments | 2.7% | 2.4% | 6.5% | 0.6% |

Source: Office of the controller of budget

**Table 8:** County Health Budget Absorption FY 2018/2019

| Details | County A | County B | County C | County D |
| --- | --- | --- | --- | --- |
| Budget absorption recurrent | 97% | 98% | 101% | 63% |
| Budget absorption development | 34% | 86% | 77% | 104% |
| Total Budget absorption | 89% | 98% | 97% | 68% |

Source: Office of the controller of budget

> *“The unutilized funds will be the opening balance for the next financial year. So your budget for the next financial year will be financed less the unutilized balance available in the account” Facility Health Manager County D*

Third, it resulted in delays in payment of staff salaries which resulted in staff demotivation that, among others manifested in absenteeism.

> *“Salaries are delayed by over a month. How should health workers continue working when they have not been paid? How should they cater for their daily needs? Once I asked a sonographer why he didn’t report to work. He told me if he left the house for work, his landlord would lock him out for failure to pay rent” Facility Manager County C*
>
> *“Salary delays have forced health workers to look for alternative sources of income. These are maintained as a cushion once salaries are paid. But they have a negative impact on the health system. The health workers’ priorities shift resulting in absenteeism thereby lowering performance. “ County Health Manager County B*

### In all the four counties, there were unclear mechanisms for priority setting during cash disbursement

It was at the discretion of treasury accountants to decide who to pay first. This had several implications on efficiency. First the payments reflected neither the health managers’ nor the patient needs. At the health facility level, the health facility managers felt demotivated. They reported that they often worked hard to raise revenue just for the money to be used in unclear ways.

> *“At both the department and county level, accountants have the ultimate say. As a director I do not know the requisitions my accounting officer has sent to treasury for payment. The accounting officer is not a medic. My priorities are not their priorities. So you end up with a very distorted payment schedule that does not address the patient needs” County health Manager County D*

Second, it introduced corruption in the payment process as suppliers were forced to lobby for payment. This reduced competitive bidding as only suppliers who were able to lobby worked with the county.

> *“There are several complaints about payments of the suppliers within the CDOH. If the challenges are experienced frequently, then pending bills accumulate. This forces people to prioritize which in turn results in lobbying. Those who are able to lobby are given first preference but there are other small suppliers who don’t have the muscles to lobby” County Finance Officer County B*

### Procurement and Supply Chain

The respondents noted that the procurement process influenced efficiency of health systems through various ways 1) tender management and competitive bidding 2) stakeholder involvement 3) Value for money 4) Timeliness of the process 5) supervision of suppliers 6) Quality of goods and services and 7) Accountability mechanisms and 8) Use of KEMSA as a single supplier.

### Respondents in three of the four counties noted that tenders were not competitively awarded

For example, in county B and D, contracts were awarded based on political patronage. In county C, contracts were awarded to companies that could pay kickbacks. This had several implications on efficiency. One, contracts were awarded to companies that were not qualified. This resulted in delivery of substandard goods and services including buildings which would be uninhabitable less than 5 years after their completion.

> *“In my opinion, county contracts are not awarded competitively rather, on political terms. As a result, most of the goods and service delivered are substandard. They build structures without toilets or running water. How can a theatre operate without running water?” Facility Health manager County D*

Second, it was felt that failure to award tenders competitively exacerbated the problem of misplaced priorities during payment. Interview respondents reported that people who had links to government officials were more likely to be paid first. This resulted in a situation where neither the county nor the health workers were interested in patient needs and experiences.

> *“Some of the suppliers are relatives to the politicians so if you are a relative or friend you’ll be paid. But the others are not paid. Whether the suppliers supply or not, we don’t get involved. When they fail to supply food, patients either buy or their relatives bring food from home. Nobody cares*.*” Sub County Health Manager County B*

Third, it reduced accountability over services and goods procured. Because the suppliers had connections, it was impossible for the health workers to condemn substandard goods or to raise issues over the quality of the goods or services. The health workers faced sanctions when they raised questions about suppliers. Besides, suppliers were still paid even after supplying substandard goods and services.

> *“According to the public procurement act, procurement should be competitive. However, the top management interferes with the process. It might look competitive on paper because they don’t want to be in problems, but it is not. If it was competitive and the contractor offers substandard services, then the county will not pay, but if they are offering substandard services and they are still paid it means there is an influence” Sub County Health Manager County B*

### Respondents noted that procurement processes were bureaucratic, lengthy and characterized by delays

The lengthy bureaucratic process influenced efficiency in several ways. First it resulted in delayed delivery of services. For example, in county C it was reported that the lengthy procurement process delayed services even when the resources were available.

> *“Delivery takes too long. This has affected service delivery. Currently, at the county referral hospital patients are buying everything, including gloves, needles for anesthesia, betadine for cleaning operation site before an emergency CS, sutures for stitching after a surgery, everything. If the patents do not have money, then we don’t do the procedure” Facility Health Manager County C*

Second, because of the long procurement process, then suppliers overpriced goods and services because they anticipated that the procurement process will consume more time and resources.

> *“Goods and services are overpriced because of the expected delay in payment. I may order in July then they’re delivered in December. Suppliers adjust for inflation and the bureaucracies of getting a tender thereby increasing the cost. “ County Health manager County C*

Thirdly, health workers in emergency situations and small health facilities were forced to contravene the procurement laws; because the process would increase the cost and timeliness of acquisition they opted for direct procurement. This direct procurement created loopholes for misappropriation of funds especially when left unmonitored.

> *“We procure small quantities so we avoid the policy of procurement protocols. What we have is little, taking it through procurement would not make sense. We do not do tendering and vetting; we just buy locally*.*” Facility Health Manager County C*

### The procurement process did not result in value for money

Value for money was compromised in two ways. First, in some instances, the counties paid for poor quality or undelivered goods. Health workers were forced to work with poor quality products which still incurred the market price for high quality goods:

> *“We have specifications that have to be met. Previously, I almost lost my job because I rejected reagents worth millions of shillings. The reagents did not meet our specifications neither did the supplier maintain cold chain during transportation as required. I was condemned but I stood my ground. The supplier returned the goods. They were to replace but I don’t know whether or not it was replaced but the supply was paid” County Health Manager County C*

Second the government paid more than the market price for commodities.

> *“It doesn’t give value for money because sometimes the quotation made for renovating a building is worth another building* .*” Sub County Health Manager County C*
>
> *“Whatever budget we have is less than what it should be. We can have a billion shillings but in terms of worth it is five hundred million. You end up doing very few things at a very high price. The budget and outcomes do not correlate” County health Manager County B*

### In two of the four counties, frontline users of goods and services were either not involved, or inadequately involved in the procurement

While the procurement process requires that frontline staff are involved at key steps of the procurement process, this did not always happen. Failure to involve the users in the procurement process led to the procurement of items that did not meet the user expectations.

> *“Neither the CDOH nor the health facility managers are consulted prior to commencing construction. We come in during inspection. Health facility managers find buildings coming up, when they question, they are told to mind their business. The building will be completed then condemned at inspection*.*” County Health Manager County B*

### Counties were required to procure from the Kenya Medical Supplies Agency (KEMSA) as their first source for medicines and medical products

KEMSA is the country’s central public procurement and supplies agency for healthcare commodities. The requirement for counties to exclusively procure from KEMSA had both positive and negative implications for efficiency. Unlike private suppliers, KEMSA sustained supplies during cash disbursement delays. This enabled continuity of service provision thereby enhancing efficiency.

> *“we delayed paying KEMSA because of the delay in the disbursement of funds from the national treasury. The CEC persuaded KEMSA to supply us and they agreed despite us owing them 19 million*.*” County Finance Manager County B*

Second, KEMSA also offered cheaper prices than some of the local suppliers thereby providing value for money.

> *“KEMSA provides value for money because they supply at the quoted price. The prices and quantities are clearly outlined. When you procure from other traders you may not realise value for money, their pricing is a bit crazy. The cost per unit pack is higher*.*” Facility health manager county D*

However, KEMSA compromised efficiency when they were unable to supply all the required medicines and supplies leading to interruptions in health service delivery.

> *“KEMSA doesn’t stock lab reagents. They cannot fully furnish our requests. They only provide grouping reagents, stool containers, and urine containers. Yet we cannot work without reagents*.*” County Health Manager County B*

### Provider Autonomy

#### Health facilities in all the four counties had either partial autonomy or no autonomy over their own source revenue

PFM rules required that all revenues generated are sent to the county revenue fund (CRF) rather than retained in health facility accounts. While both the respondents and the document review indicated that there were existing legal provisions for health facilities to retain their revenue, most counties were unwilling to explore this.

> *“All revenue generated by the CDoH goes back to the county. For a long time finance was quoting the PFM act insisting that they require a law for CdoH to use its revenue at source. We finally obliged and created the law that was passed by the assembly. But, it has not been operationalized to date” County Health Manager County B*
>
> *“Patients are dissatisfied with our services because we do not have funds at the facility. Ideally the money should go to the CRF, but it should be sent back to the health facilities in less than fifteen days but that has not happened since October*.*” County Health Manager County B*

Lack of financial autonomy, whether partial or complete had several implications on efficiency. First, health facilities had constrained access to resources. As a result, the health system was unable to adequately deliver health services.

> *“The County government used to purchase supplies for lab and radiology. But they no longer purchase. We buy all these supplies. They consume a lot of our money. We charge patients for laboratory and radiology services. The county collects all the money. Then we are left without resources to replenish supplies” Facility Health Manager County A*

Second, the process for health facilities to access funds within the CRF was long and bureaucratic resulting in delays in provision of health services.

> *“In case of an emergency, you might lose a patient because you lack essential commodities. And this happens frequently. That is the downfall of a health system that not properly financed” Sub County Health Manager County B*

Third, limited autonomy compromised the link between financial and performance accountability. Health facilities were unable to effectively evaluate their performance as they were unaware of the full extent of their expenditure.

> *“You see there is what the county spends on the facility and then there is what I spend*. *My expenditure comes from NHIF reimbursement. This I can evaluate what worked and what didn’t work, and improve on subsequent expenditure. But it’s hard to evaluate what the county probably spent on us*.*” Facility Health Manager County A*

#### In three of the four counties the health department control over the procurement process was limited

Power over what should be procured, and when, was with the county treasury. This compromised efficiency by limiting service delivery and misaligning procured commodities with needs at the health facility level.

> *“Finance issued a tender to purchase an anesthetic machine. The supplier was unable to get the specifications requested in the tender. He came to consult us, only to realize, the user, the anesthetist wasn’t consulted on the specifications. Going forward, we want the users to decide what is to be bought. This decision should not be left to anyone who has access to the money “ County Health Manager County B*

### Financial Management System

County governments used the Integrated financial management system (IFMIS) to process all government payments. The use of this system was rolled out in 2018 to enhance accountability of funds. This system has had several effects on the efficiency of county health system. Respondents noted that IFMIS has traceability and hence improved financial accountability. Respondents also felt that IFMIS also improved efficiency by digitising PFM processes. However, this was limited by the requirement that counties provide printed documents to the national government for approval.

#### Access to the IFMIS was limited to a few people, and even amongst the few people, there were varying levels of access

As a result, it has limited access to information thereby exposing health funds to misuse and limiting the departments autonomy over their resources.

> *“the chief officer finance has more powers in the system, he can reallocate budgets between departments. There are instances when we request to execute part of our budget, but the request is declined because the allocation is exhausted. Yet the CDOH did not spend the resources. My chief officer cannot claim with certainty that we have resources on a certain vote. IFMIS payment powers should be devolved to the departmental level*.*” County Health Manager County B*

## DISCUSSION

We examined the relationship between the budget execution process and the efficiency of county health systems in Kenya. We found that each dimension of the budget execution process has potential effects on health system efficiency. For the county health departments in Kenya, the budget execution process defined the implementation of their workplans and almost all service delivery activities.

We set out to explore the relationship between budget execution processes and efficiency in two efficient and two inefficient counties. Challenges within the PFM system were generally cross cutting, with no clear distinction between the efficient and inefficient counties. However, one county stood out. County A, one of the efficient counties had more credible budgets, more efficient cash disbursement processes, partial provider autonomy and more efficient procurement systems. This perhaps provides some evidence that effective PFM processes enhance the efficiency of health systems. However, while county B was ranked as efficient, we found that it shared similar PFM challenges with the inefficient counties. This mixed finding could be because the nature of PFM practices documented are perverse in Kenyan counties, with differences in degrees across countries that are difficult to tease out using a qualitative approach. It could also be because the counties that were ranked as efficient by the quantitative analysis by being on the efficiency frontier are inefficient in absolute terms, even though they are relatively more efficient than the counties that are at a distance from the efficiency frontier. This not with-standing, the study found that county budget execution challenges could potentially influence the efficiency of county health system in several ways.

First, some budget execution practices are likely to compromise the input mix of county health systems with negative impacts on the capacity of county health systems to deliver healthcare services and consequently health outcomes. We found that county health budgets were hardly credible, characterised by the failure of county governments to honour budget allocations to county health departments, and delays in the disbursement of funds. These delays and non-disbursement of funds thus constrained the resources available to county health departments and reduced the county health departments budget absorptive capacity. Delays in procurement also impacted negatively on service delivery. Studies from other settings have documented how the lack of credibility of budgets limits efficiency by disrupting service delivery and impairing manager’s ability to implement health system plans (Piatti-Fünfkirchen *et al*., 2021). It has also been shown by other studies that cash disbursement delays may lead to rushed spending and poor budget absorption when funds are availed at the end of the financial year (Glenngård and Maina, 2007; Abekah-Nkrumah, Dinklo and Abor, 2009; Piatti-Fünfkirchen *et al*., 2021).

Delays in funds disbursement and procurement were also because of the limited financial and managerial autonomy of county health departments and health facilities. Other studies in Kenya have demonstrated that limited provider autonomy over managerial and financial roles hinder service delivery (Barasa *et al*., 2017; de Geyndt, 2017). The main reason for the reduced autonomy of county health departments and health facilities is the requirement by the PFM laws for all funds to be managed centrally from the CRF account (Government of the Republic of Kenya, 2012). It has however been observed that it is possible to provide autonomy to county health departments and health facilities under existing PFM laws suggesting the potential that the lack of autonomy is an intentional misinterpretation of PFM laws informed by county leaders interest to control resources centrally. This study found that autonomy was additionally compromised by several practices including limited access to the financial management information systems. Similar findings were reported in Tanzania where investment in a financial management information system that could be implemented to the lowest planning unit enhanced reporting and accountability thereby enhancing health system efficiency (Piatti-Fünfkirchen and Schneider, 2018b).

Second, inefficiencies could also arise due to misalignment between county health needs and the use of resources. We found that actual county department of health expenditures deviated from approved budgets. Further, we found that cash disbursements by county treasury accountants used unclear considerations resulting in the prioritization of expenditures on activities that were not aligned with county health department priorities. In Kenya, one study reported reallocation of health system resources to fund the governor’s promises (Waithaka *et al*., 2018). In Democratic Republic of Congo (DRC), health funds were used to finance administrative activities in the office of the governor (le Gargasson *et al*., 2014). Rent seeking and political patronage of procurement processes also contributed to the misalignment of payments and county department of health priorities. Misalignment also resulted from the inadequate involvement of frontline health workers in the procurement process for goods and services, and the reduced financial and managerial autonomy of county health departments and health facilities.

Third, county health system efficiency could be negatively impacted by reduced staff motivation and productivity. We found that delays in funds disbursements resulted in late payment of staff salaries leading to demotivation. Health facilities managers were also demotivated by the lack of alignment between their stated priorities as articulated in approved budgets and plans and the county treasury accountants revealed priorities by disbursing funds for specific activities. In Yemen and Ghana, cash disbursement delays led to halting of key activities and delays in salaries that demotivated employees (Asante, Zwi and Ho, 2006; Elgazzar, 2011).

Fourth, procurement inefficiencies, that included the procurement of substandard goods and services, and the inflation of procurement prices by supplies of goods and services could negatively impact county health system efficiency. We found that county contracts were sometimes not competitively awarded, and instead influenced by political patronage and bribes, sometimes resulting in substandard goods and services. Also, suppliers of goods and services to counties often inflated procurement prices because of anticipated delays in payments by county governments. Similar findings are reported in Czech republic and South Africa where poor procurement practices resulted in inefficiencies (Global Access Partners, 2016; Munzhedzi, 2016; Transparency International Česká republika, 2016).

Lastly, inefficiencies could arise from compromised county accountability for finances and performance. Staff were not empowered to reject sub-standard goods and services because of the political patronage enjoyed by suppliers. Accountability for performance was also compromised by the reduced financial and managerial autonomy of county departments of health and managers. Further, delays or non-disbursement of funds, and the misalignment of county department of health’s plans and actual expenditures meant that county managers had reduced legitimacy to hold staff accountable for performance. In Tanzania and Zambia, reduced managerial autonomy limited their accountability over performance (Piatti-Fünfkirchen and Schneider, 2018a).

This study had several limitations. First, we only sampled 4 out of the 47 counties in Kenya. Budget execution processes within counties are diverse. Besides, given the semi – autonomous nature of counties in Kenya, their sources of revenue, and laws are equally diverse. Second, we collected budget data from multiple documents at both county and national levels. Besides the data was extracted at different time points. Where the data conflicted, we used the most complete data source.

Despite the limitations, county governments need to improve their budget execution processes to improve the health system efficiency. First, counties should make realistic own source revenue projections. This will ensure that budgets ceilings are realistic and set the ground for a credible budget. Second, counties should strive to meet conditions attached to conditional grants to enhance credibility of conditional grants. Third, the CDOH should be involved in prioritizing payments to ensure that payments reflect the needs of the departments. Fourth, the government should improve timeliness in the procurement process including timely payments, this will reduce the cost of goods and services and encourage competitiveness as more suppliers will be motivated to participate in the procurement process. Fifth, county governments should implement mechanisms to ensure that providers have more managerial and financial autonomy. Sixth, the government should increase provider autonomy over both their financial and managerial functions. This will possibly increase transparency in the budget execution process, align budget execution to health systems needs and increase health facility managers accountability for performance. Finally, the government should roll out IFMIS to the lowest planning unit. This will increase transparency and enhance accountability over resources thereby enhancing efficiency.

## Conclusion

In conclusion, all the five aspects of budget execution influence health system efficiency directly or by influencing the budget formulation evaluation processes. While a well formulated budget is a good starting point for efficiency within the health system, the budget should be well implemented to realize the desired outputs and outcomes.

## Data Availability

All data produced in the present study are available upon reasonable request to the authors

